# Assessing the benefits and risks to mothers and offspring of continuing treatment for maternal hypertension and hypothyroidism: an observational cohort study in the UK Clinical Practice Research Datalink

**DOI:** 10.1101/2024.08.09.24311727

**Authors:** Ciarrah-Jane S Barry, Venexia M Walker, Christy Burden, George Davey Smith, Neil M Davies

## Abstract

**AJOG at a glance:** *A. Why was this study conducted?*

This study aimed to evaluate the risks and benefits of discontinuing maternal prescriptions for chronic hypertension and hypothyroidism during pregnancy.

*B. What are the key findings?*

Discontinuing beta-blockers and thyroid hormones was associated with some improved maternal outcomes, such as a reduced risk of miscarriage. We found limited evidence of an association between the discontinuation of multiple drugs subclasses such as renin-angiotensin system drugs and differential risk of maternal or neonatal outcomes.

*C. What does this study add to what is already known?*

This study provides observational evidence that discontinuing certain medications may not be strongly associated with differential risk of maternal or neonatal outcomes and highlights the need for further research due to limitations such as confounding by indication and potential data misclassification.

Exclusion of pregnant women from clinical trials, due to ethical concerns, has limited evidence on medication safety during pregnancy, resulting in conservative guidance. Yet, rising prevalence of chronic conditions in reproductive-age women has increased medication use. This study evaluates risks and benefits of discontinuing drug prescriptions for chronic hypertension, and hypothyroidism during pregnancy using linked primary care records from a longitudinal intergenerational database.

Using UK Clinical Practice Research Datalink (CPRD) GOLD, we conducted multivariable regression models, adjusted for covariates, to assess maternal treatment discontinuation on various outcomes.

Cohorts of 3,232 and 3,334 pregnancies with chronic hypertension and hypothyroidism respectively were derived from the CPRD. Discontinuing vasodilator antihypertensive drugs for hypertension was associated with increased gestational age (mean difference: 3.98 weeks, 95% CI: 1.61, 6.35). Estimated associations between other antihypertensives (calcium-channel blockers, diuretics or renin-angiotensin system drugs) and any study outcome crossed the null. Discontinuing thyroid hormones for hypothyroidism were associated with reduced the odds of miscarriage (OR: 0.29, 95% CI: 0.15, 0.54) and increased gestational age (mean difference 1.84 weeks, 95% CI: 0.12, 3.57). Results were robust in sensitivity analysis.

This study reports a reassuring lack of association between many drug subclasses and adverse offspring outcomes. This evidence of potential risk associated with treatment discontinuation may guide clinical decision-making for treating chronic hypertension and hypothyroidism during pregnancy in similar populations.

## Introduction

Pregnant women are typically excluded from pharmaceutical trials (1–3), unless the drug under study only applies to a pregnant population (2,4). Further, women may be reluctant to participate in clinical trials due to the potential harm thought to be caused to the baby by treatments (5). Observational studies are therefore one of the primary sources of evidence about the intrauterine effects of maternal medication use (6). Thus, medications are often prescribed in pregnancy with limited evidence about the effects on the mother and child. Many mothers need to continue their treatments started before pregnancy, as active chronic disease can be associated with adverse pregnancy outcomes.

Despite the limited evidence, the use of prescriptive medication during pregnancy has increased globally, as the number of pre-existing chronic medical conditions in the population of reproductive age has risen (7,8). A recent survey suggested hypertension affects 3% of women in England aged 16-44, while hypothyroidism has a prevalence of 3.6% in the UK, and is more commonly diagnosed within women (9–11). These conditions can be worsened by the physiologic changes throughout pregnancy, such as differing insulin resistance, fluctuations in blood pressure, and circulating concentration of thyroxine-binding globulin (TBG) (12–14). Left uncontrolled and untreated, these conditions are established risk factors for a range of adverse maternal and neonatal outcomes, such as macrosomia, postpartum haemorrhage, premature birth and developmental abnormalities and perinatal death (15,16).

Individual case reports, post-marketing surveillance data, retrospective cohort studies and animal trials within the literature have provided evidence of potential adverse neonatal outcomes. Examples of this include intrauterine growth restriction, neonatal hyper- or hypoglycaemia and neonatal death associated with exposure to some of these medications throughout gestation, such as some beta-adrenoreceptor blocking drugs (17,18).

Current guidelines and advice recommend that treatments are only prescribed when the benefit of treatment is thought to outweigh the risks of unknown drug exposure to the fetus (6,19). This is assessed on a patient-by-patient basis by the clinician and can be an extremely challenging judgement given the available evidence of safety during pregnancy (20).

We aimed to assess the risks and benefits associated with discontinuing maternal drug prescriptions for the treatment of chronic hypertension and hypothyroidism in both mothers and offspring, using linked primary care records within a longitudinal intergenerational database to add to the body of evidence for clinical decision-making with medication use in pregnancy.

## Methods

### Data source

The Clinical Practice Research Datalink (CPRD) GOLD is a longitudinal electronic healthcare record (EHR) database in the United Kingdom (UK) containing approximately 11 million anonymised patients across 674 primary care practices (21). These patient-level primary care records contain routinely collected data, including diagnoses, prescriptions, and demographic characteristics, from participating general practice (GP) surgeries. The GP is the primary point of contact for non-emergency health-related issues within the UK. Relative to the population in the 2011 census, the CPRD is broadly representative for age, ethnicity and sex, with the majority of the population registered with a GP (21–23). The data entered is subject to extensive quality control and validation (2).

Patients’ primary care data was linked to Hospital Episode Statistics (HES) Admitted Patient Care, death registration data from the Office for National Statistics (ONS), and the Index of Multiple Deprivation (IMD) 2011. Linkages are available for approximately 58% of practices within the CPRD (21). Linkages are only available for practices in England (391 of 674 practices).

Medical history and diagnosis information are coded using Read codes, and medical codes are codes used for medical terms used by the general practitioner (GP) within primary care files (25,26). Items that may be prescribed are coded by product code (e.g., codes used to record treatment by the GP) (26). Within HES England, surgical procedures, operations and interventions are coded using the Classification of Interventions and Procedures (OPCS-4) codes (27). Diagnoses within HES are coded according to the UK International Classification of Diseases 10th Revision (ICD 10) codes (27).

The CPRD has also developed an algorithm to link likely mother-offspring pairs within the primary care data, which forms the mother-baby linkage dataset. A further algorithm was developed in 2019 to generate a Pregnancy register linkage to characterise and identify pregnancies within CPRD GOLD systematically (28). Data for this project was extracted from the June 2021 Primary care build (ISAC protocol number 21_000362).

### Study population

All mothers were women, aged between 11-49 years old and on the Pregnancy register between April 1997-March 2021 inclusive (29,30). Mothers were diagnosed with a condition of interest at least a year before the start of pregnancy and received at least two relevant prescriptions of the same BNF drug subclass for their condition in the six months before the estimated start of pregnancy. Within the pregnancy register, pregnancy start dates were estimated from the estimated delivery date, estimated date of confinement (the predicted date of expected birth calculated from last menstrual period and estimated gestational age), last menstrual period, gestational age at birth, gestational age from the antenatal record or imputed from the earliest antenatal entry in the mother’s primary care record (28). All mothers were required to have at least 12 months before the start of pregnancy “up-to-standard” practice level data, as defined by the CPRD.

Lists of medical codes were developed, refined, and clinically validated to identify mothers diagnosed with the relevant condition of interest. Lists of product codes were developed, refined, and clinically validated to identify relevant prescriptions within maternal primary care data.

### Exposures

We investigated the impact of treatment discontinuation on maternal and offspring outcomes. Maternal exposure was defined as receiving a prescription for a drug in the same drug subclass as before pregnancy, after the start of pregnancy. We performed our analysis at the drug subclass level, as indicated by the British National Formulary (BNF).

### Outcomes

Maternal outcomes were defined using a combination of the pregnancy register, HES and primary care data, see Supplementary Table 1. These were the method of delivery (vaginal, forceps/vacuum, other delivery, unspecified, caesarean, breech), the incidence of urinary tract infection (UTI), postpartum haemorrhage, poor hypertensive control during pregnancy, miscarriage, poor thyroid control during pregnancy and gestational diabetes mellitus.

For linked mother-offspring pairings in which the offspring was designated a patient identifier, neonatal outcomes were defined via the Pregnancy register, both maternal and neonatal HES and primary care data. For pregnancies that did not result in a linked mother-offspring pair, neonatal outcomes were identified via the Pregnancy register, maternal primary care and HES data. These were stillbirth, severe preterm birth (defined as a birth between 22 and 37 weeks of gestation), preterm birth (defined as a birth after 37 but before 40 weeks of gestation), gestational age and congenital malformation(s).

### Covariates

Sociodemographic differences and comorbidities may influence prescribing patterns. We accounted for this through the inclusion of the following covariates: historical incidence of postpartum haemorrhage, historical miscarriage, historical stillbirth, historical preterm birth, multiple birth, smoking status prior to pregnancy (current/former/never), a binary measure indicating ‘teetotal’ prior to pregnancy, region (as defined by CPRD), maternal age, relationship status, maternal body mass index (BMI, kg/m^2^) up to two years prior to the start of pregnancy, maternal ethnicity, parity and sex of baby. See Supplementary Notes and Supplementary Table 1 for additional details of variable derivation. To account for the potential effects of additional medical conditions, the Cambridge Multimorbidity Index (CMM) was calculated per pregnancy. The CMM is a weighted score that may be calculated to quantify multiple patient health conditions likely contributing to poor health outcomes (31). We also included the frequency of primary care visits with any staff member in the year prior to the start of pregnancy to account for the potentially higher ascertainment of outcomes in those with more contact and calendar year of delivery to account for temporal differences in prescribing patterns.

To maximise sample size, a hierarchal search was applied to derive study variables from primary care, HES and the Pregnancy register (Supplementary Figure 1).

### Missing data

Separate imputations were run based on the length of gestation as only pregnancies with gestational age over 22 weeks were viable for particular outcomes. For pregnancies with any gestational age, we imputed relationship status, ethnicity, maternal BMI, parity and sex of baby. For pregnancies that had gestational age of at least 22 weeks, we additionally included delivery method. For the covariates and outcomes based on a diagnosis in patient primary care records, the absence of a code and no other evidence within the patient’s medical record was assumed to reflect the absence of the event for each pregnancy. We could not discern missing data for these variables.

The Pregnancy register was derived using primary care data. To recover information about ‘unknown’ pregnancy outcomes and differentiate between conflicting pregnancy episodes, we applied a proxy pregnancy-register algorithm to linked HES data (32).

### Statistical analysis

Multivariable linear and logistic regression models were run for continuous and binary outcomes respectively, adjusting for the covariates to control for measured confounding. Robust standard errors were calculated. Clustering over patient ID was necessary to account for potential trends in prescribing at the patient and practice level, and also the inclusion of multiple pregnancies per maternal patient identifier. Mean differences and 95% confidence intervals (CI) were calculated for continuous outcomes. Odds ratios and corresponding 95% CIs were calculated for binary outcomes.

All analyses were performed using R version 4.1.1. The pre-specified protocol was shared before the commencement of analysis (33). Analysis code and code lists are available online: https://github.com/Ciarrah/CPRD.

### Sensitivity analysis

Propensity score regression was performed as a sensitivity analysis. A propensity score was calculated from known covariates and included in the regression model to enable the calculation of the treatment effect whilst controlling for all covariates (34,35). Assuming the model is correctly specified, this method controls for confounding, increases estimate precision and reduces bias (36,37). We must assume the probability of treatment is not influenced by potential outcomes or unobserved variables. We visually assessed the covariate balance between treatment groups to determine whether the covariate balance was sufficient by plotting the propensity score.

We then performed propensity score regression to assess whether the addition of the propensity score improved our estimates. Further, we applied a trimming threshold of 1% on both tails and performed propensity score regression in this subset to assess the sensitivity of discontinuation to the exclusion of patients who would receive their allocation irrespective of all measured factors (38).

### Deviations from protocol

In our pre-specified protocol, we intended to include Apgar score, offspring birth weight, maternal death and neonatal death as outcomes (33). Due to high levels of missing data, these outcomes were removed. Additionally, before data acquisition, we proposed investigating maternal pre-existing diabetes and hyperthyroidism. There was insufficient variation in the therapeutic decision-making to include pre-existing diabetes, and insufficient sample size to include hyperthyroidism.

Initially, within the propensity score analysis of the hypothyroid cohort, we included a comprehensive set of all covariates, as they are theoretically associated with treatment choice and outcome. We reviewed the standard errors of the model and reduced the model to the set of variables that had high uncertainty to ensure propensity score overlap between treatment groups. This meant the individuals in both treatment arms were comparable, and we achieved covariate balance, reducing bias in the estimated treatment effect.

We intended to perform a matched sibling analysis of sibling pairs whose mothers had received discordant prescribing patterns for their respective pregnancies. However, we were unable to implement this analysis as we had insufficient sibling pairs.

## Results

From the CPRD database, we identified 3,232 and 3,334 mothers for the cohorts of antihypertensives and hypothyroid treatments respectively, Figure 1.

**Figure 1:**
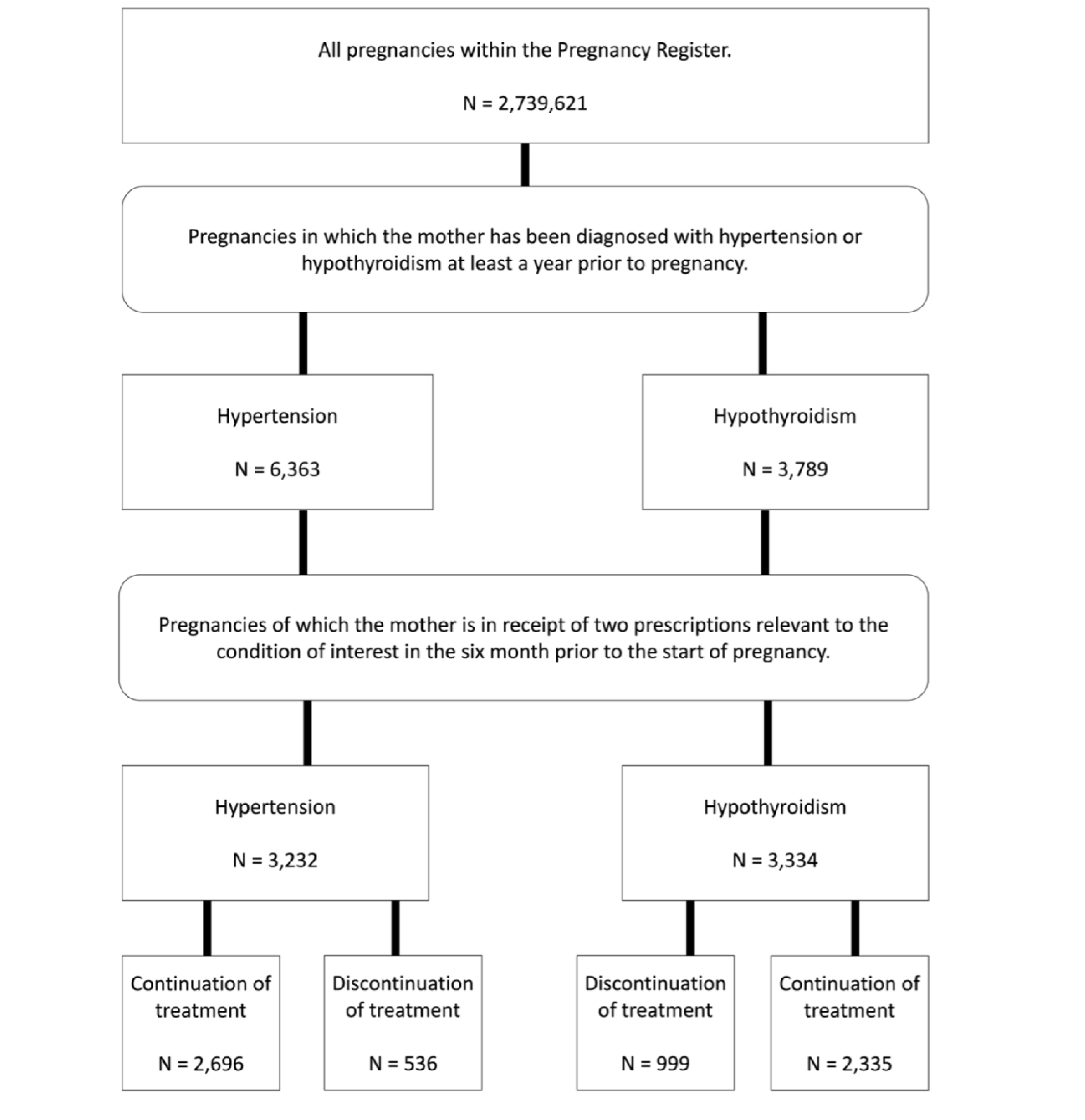
A flowchart demonstrating cohort inclusion criteria.

**Figure 2:**
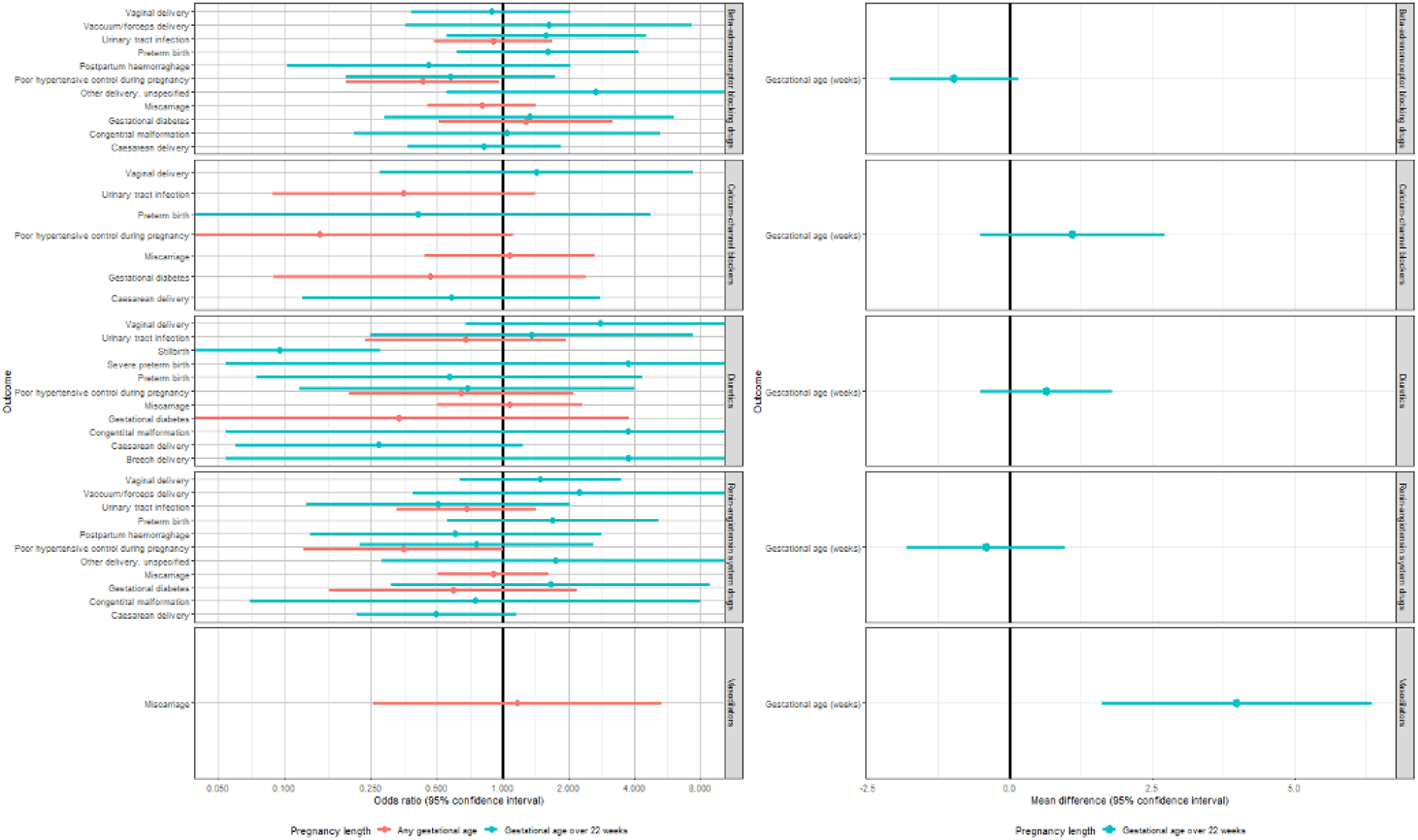
A forest plot demonstrating the estimated association between maternal discontinuation of antihypertensive treatment and maternal and neonatal outcomes using multivariable regression. An odds ratio (OR) and 95% confidence interval (CI) has been calculated for binary outcomes (left), the mean difference and 95% CI has been calculated for continuous measures.

**Figure 3:**
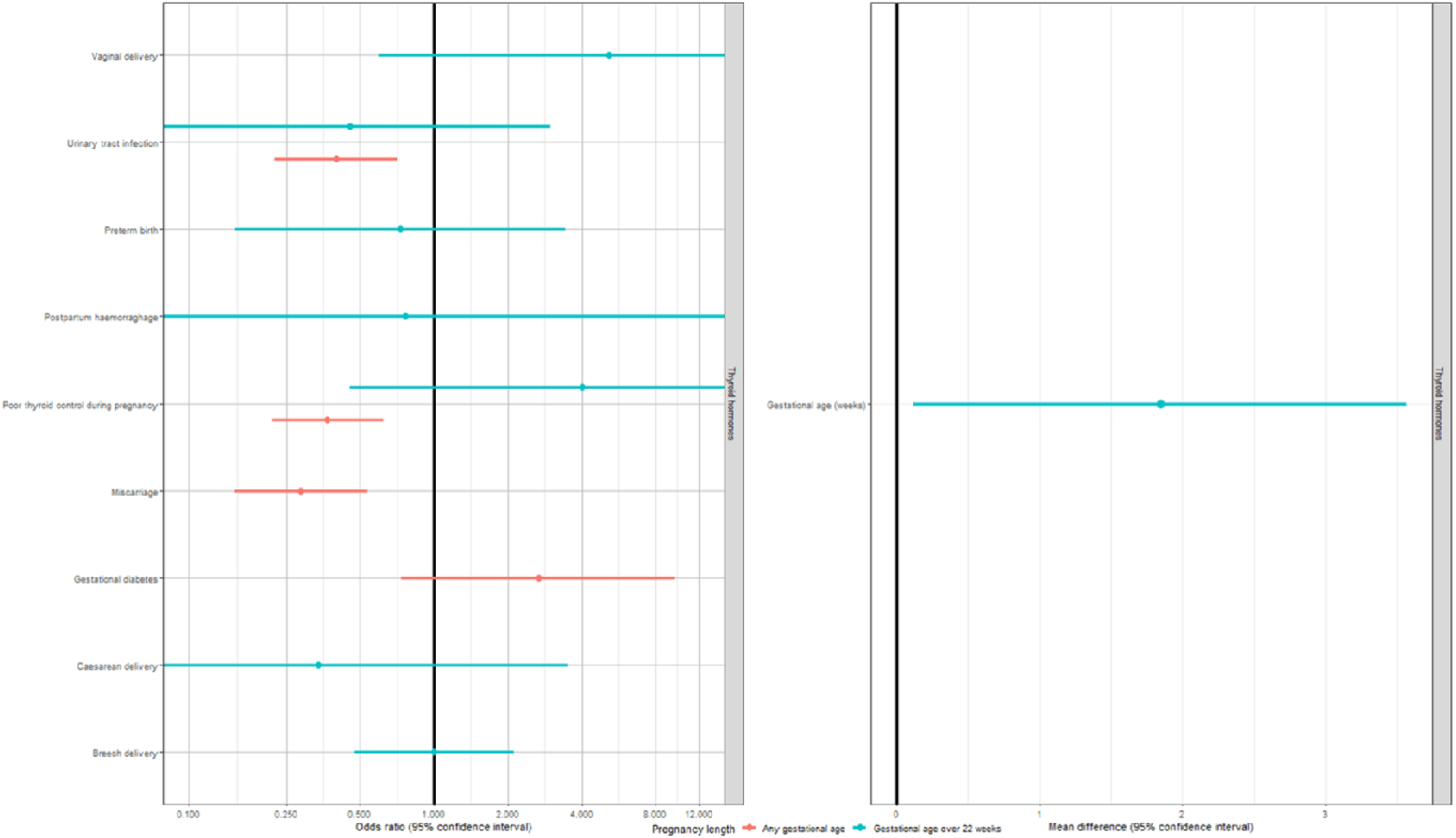
A forest plot demonstrating the estimated association between maternal discontinuation of treatment for hypothyroidism using multivariable regression. An odds ratio (OR) and 95% confidence interval (CI) has been calculated for binary outcomes (left), the mean difference and 95% CI has been calculated for continuous measures.

Of the included patients with hypertension or hypothyroidism, 197 and 25 had complete covariate information respectively. As the majority of the cohorts had some missing information, we performed multiple imputation. Patient characteristics for those included in our cohorts are presented in Table 1.

**Table 1:**
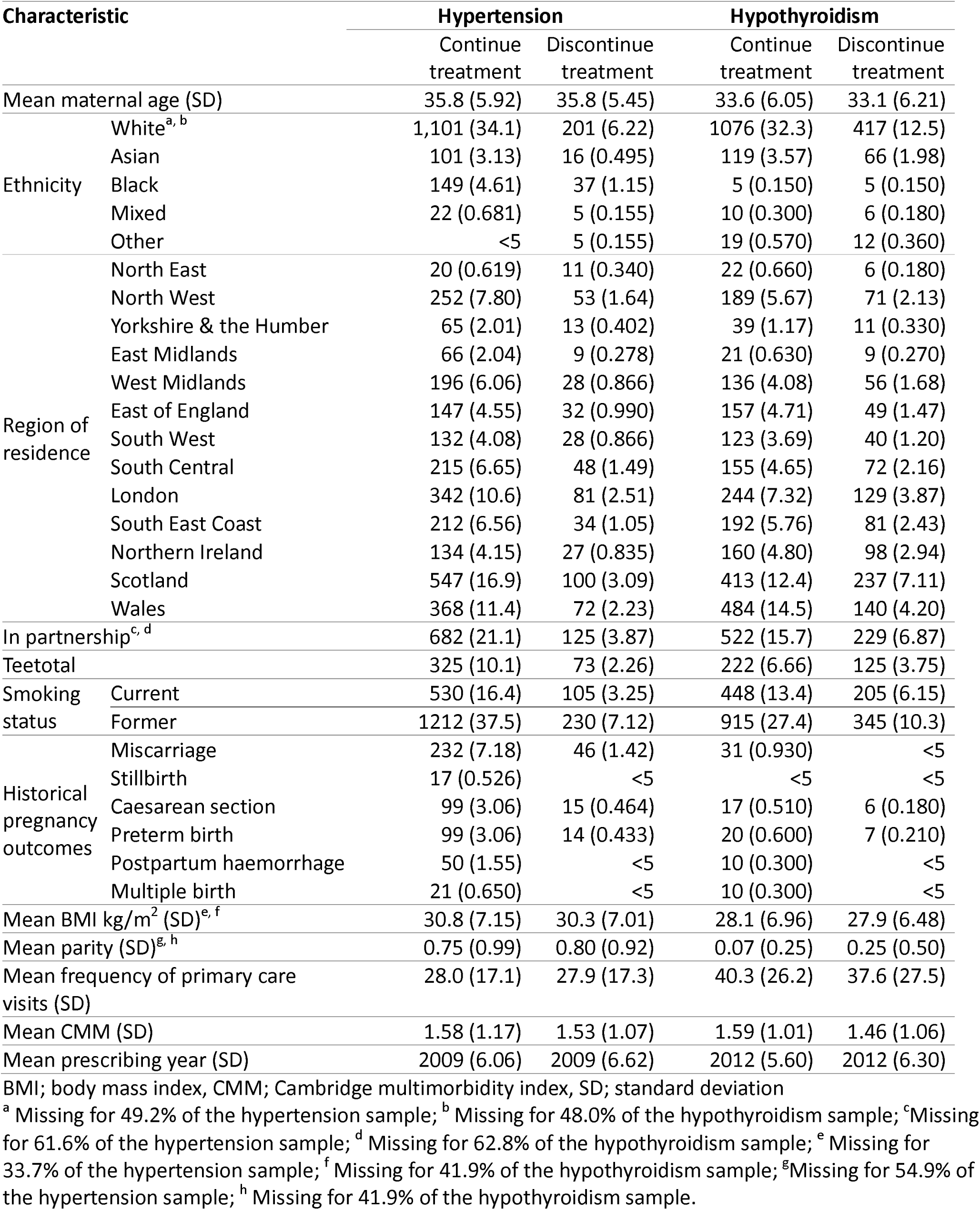
Descriptive pregnancy characteristics of the study sample.

Characteristics of pregnancies that continued or discontinued treatment were similarly distributed across measured covariates, meaning our exposed and unexposed participants did not systematically differ.

The number of pregnancies exposed to each drug subclass is demonstrated in Table 2.

**Table 2:**
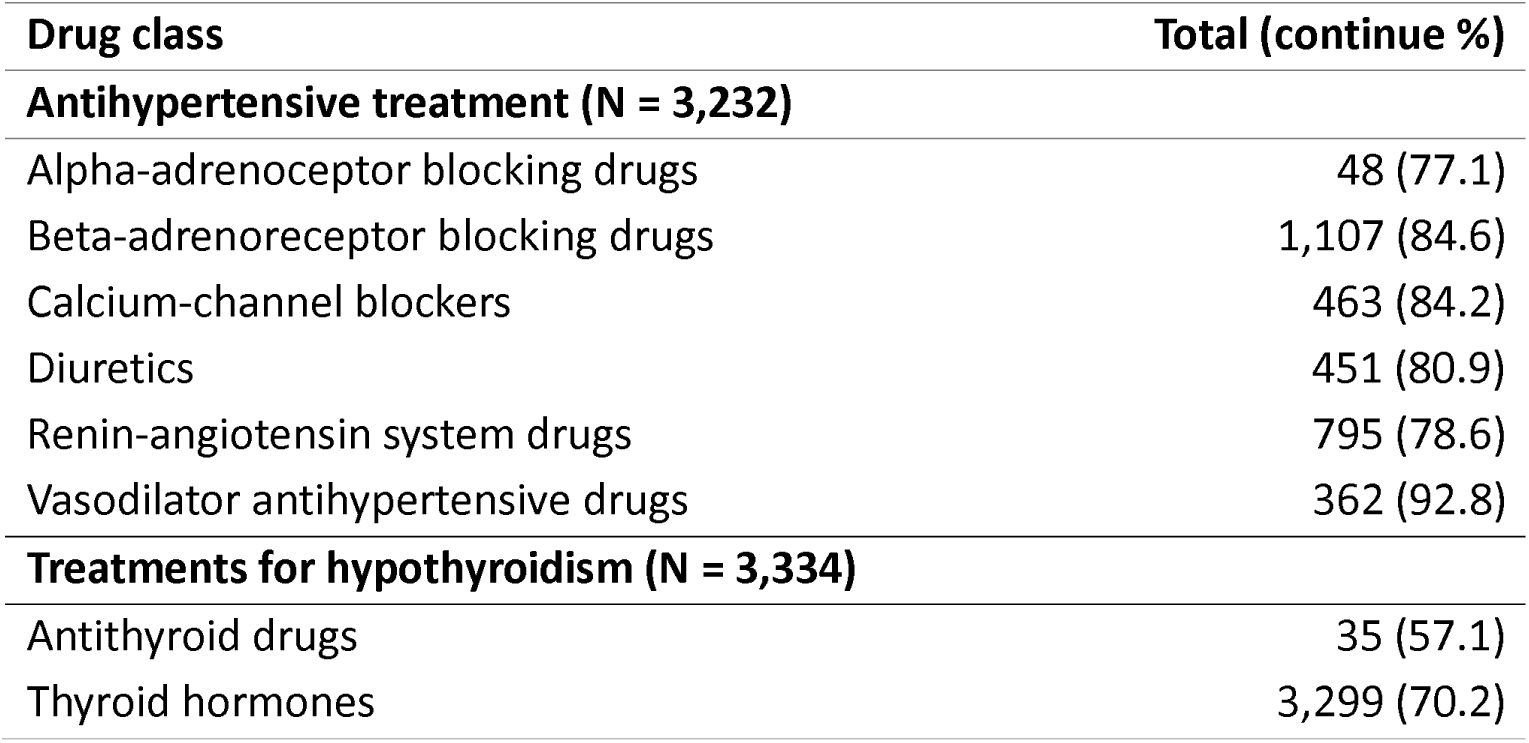
Counts of pregnancy treatment exposure by drug subclass.

| Drug class | Total (continue %) |
| --- | --- |
| **Antihypertensive treatment (N = 3,232)** |  |
| Alpha-adrenoceptor blocking drugs | 48 (77.1) |
| Beta-adrenoreceptor blocking drugs | 1,107 (84.6) |
| Calcium-channel blockers | 463 (84.2) |
| Diuretics | 451 (80.9) |
| Renin-angiotensin system drugs | 795 (78.6) |
| Vasodilator antihypertensive drugs | 362 (92.8) |
| **Treatments for hypothyroidism (N = 3,334)** |  |
| Antithyroid drugs | 35 (57.1) |
| Thyroid hormones | 3,299 (70.2) |

### Antihypertensives

Discontinuation of a prescription for beta-adrenoreceptor blocking drugs was associated with an odds ratio (OR) of 0.42 (95% confidence interval (CI): 0.18, 0.96) for poor hypertensive control during pregnancy across all gestational ages. For pregnancies over 22 weeks, the estimate was less precise, with an OR of 0.56 (95% CI: 0.19, 1.74). Vasodilator antihypertensive drugs were the only subclass for which discontinuation of prescription was associated with an increase in gestational age, with an estimated mean difference of 3.98 weeks (95% CI: 1.61, 6.35). All estimated associations for the discontinuation of diuretics, renin-angiotensin system drugs and calcium-channels blockers with any study outcome crossed the null, for example, discontinuation of diuretics was estimated to have an OR of 1.12 (95% CI: 0.50, 2.49).

### Hypothyroidism

Across any gestational age, for patients with hypothyroidism, discontinuation of thyroid hormones was estimated to have OR of 0.40 (95% CI: 0.22, 0.71) for urinary tract infections (UTI) and OR of 0.37 (95% CI: 0.22, 0.62) for poor thyroid control during pregnancy. Additionally, discontinuation of treatment reduced the incidence of miscarriage (OR: 0.29, 95% CI: 0.15, 0.54). We did not find differential risks between the discontinuation of prescription for thyroid hormones for hypothyroidism and any method of delivery related outcome. For example, mothers who discontinued treatment had an estimated OR for breech birth of 1.00 (95% CI: 0.47, 2.11). The discontinuation of prescription for thyroid hormones was We could not estimate the association between discontinuation of prescription for antithyroid drugs for hypothyroidism and any outcome due to an insufficient number of mothers being prescribed this subclass during pregnancy.

### Sensitivity analysis

#### Covariate balance

Covariate balance for measured variables varied according to the condition and drug subclass under slightly higher propensity scores than pregnancies in which treatment was continued for both hypertension and hypothyroidism.

#### Propensity score analysis

Results were consistent between multivariable and propensity score regression analyses, Supplementary figures 4 and 5.

#### Trimmed propensity score analysis

We calculated propensity score-adjusted effect estimates excluding pregnancies with the top and bottom 1% of propensity scores, Supplementary figures 6 and 7. The decreased risk of stillbirth associated with diuretic discontinuation attenuated towards the null, yet confidence intervals still overlapped (untrimmed OR: 0.16, 95% CI: 0.07, 0.40; trimmed OR: 0.19, 95% CI: 0.10, 0.39). Here, the impact of vasodilator antihypertensive treatment discontinuation on gestational age was unable to be assessed as all pregnancies in which treatment was discontinued were trimmed from analyses. Results were consistent for hypothyroid treatment and all available outcomes.

## Discussion

### Principle findings

In this study, we examined the associated risks and benefits of discontinuation of maternal prescriptive drug use for the treatment of maternal chronic hypertension and hypothyroidism.

Discontinued prescriptions for some conditions and drug subclasses were associated with improved maternal outcomes. For example, discontinued prescription of beta-adrenoreceptor-blocking drugs for hypertension during pregnancy was associated with a reduced likelihood of poor maternal hypertensive control. Additionally, discontinuation of thyroid hormones was associated with a reduced incidence of poor maternal thyroid control. We found limited evidence to support an association between discontinuation of prescribing calcium-channel blockers or renin-angiotensin system drugs for hypertension and any study outcome. Our propensity score sensitivity analyses demonstrated little difference in covariate balance.

Results also remained consistent after the exclusion of cases with propensity scores at the upper and lower extremes. This suggests main analysis findings are unlikely to be driven by these cases. We note some variables were dropped from the trimmed analysis due to low counts.

### Results in the context of what is known

Labetalol, a beta-adrenoceptor blocking drug, and nifedipine, a calcium channel blocker, are typically prescribed during pregnancy (39–41). For the outcomes available within our analysis, we found limited evidence of an association between discontinuation of treatment and differential maternal or neonatal outcomes. UK guidance specifies diuretics and ACE-inhibitors are not routinely recommended due to the risk of altered uteroplacental blood flow (42). An observational study in Denmark and Scotland estimated maternal diuretics users to have increased risk of preterm delivery (43). The authors suggested this result may be due to confounding by indication, highlighting a diabetes prevalence of 10.3% within the Danish sample. Our study, with 11% prevalence of diabetes, did not demonstrate that discontinuation of a diuretic prescription was associated with gestational age or preterm birth risk, in agreement with a Cochrane randomised controlled trial review (44). Renin-angiotensin system drugs are typically avoided during pregnancy due to established adverse neonatal outcomes (45,46). Yet use is highly prevalent in non-pregnant individuals therefore unplanned pregnancies may experience unintended fetal exposure (47). Our results suggest that an OR associated with discontinuation and miscarriage larger than 1.62 is unlikely.

Vasodilator antihypertensive drugs, such as hydralazine, may be prescribed during pregnancy if the risk of uncontrolled maternal hypertension is high (48,49). A meta-analysis of hydralazine versus other antihypertensives found no increased risk of prematurity within pregnant hydralazine users (49). We found discontinuation of vasodilator antihypertensive drugs was associated with an increase in gestational age. Poorly controlled chronic hypertension during pregnancy may increase the risk of prematurity (50). Confounding by indication may have impacted our estimate as patients with less severe hypertension may discontinue treatment. We were unable to discern whether this association was due to maternal hypertension effects on the offspring, or treatment exposure.

Levothyroxine (thyroid hormone) is predominantly prescribed for chronic hypothyroidism during pregnancy. The drug safety profile is relatively established within pregnancy (51–53). Generally, our findings suggest that treatment discontinuation is unlikely to be associated with greatly increased odds of harmful fetomaternal outcomes. Discontinuation of thyroid hormone prescription was associated with reduced likelihood of miscarriage, potentially indicating neonatal exposure to thyroid hormones is more detrimental than uncontrolled maternal TSH. This contrasts with the literature, in which a reduction in miscarriage rate is typically found with levothyroxine treatment (51,54). Again, we note mothers that discontinue treatment may have less severe hypothyroidism so estimates may reflect maternal hypothyroid related effects on the offspring. UTI are a common complication during pregnancy and is associated with a range of adverse pregnancy outcomes (55,56). We found that thyroid hormone treatment discontinuation was associated with reduced odds of UTI. This may contribute towards the overall reduction in likelihood of UTI related adverse pregnancy outcomes, or residual confounding e.g. by maternal health.

### Strengths and limitations

There are numerous strengths of the CPRD. First, primary data can be linked to a wide range of external datasets which increases the span of possible analyses and enables inclusion of a greater range of variables such as socioeconomic position. Second, it enables the determination of drug safety information through observational studies in circumstances in which an RCT is unethical or infeasible. Third, study findings are typically generalisable to the UK, and other populations of similar demographics with public national healthcare systems, as the CPRD is thought to be widely representative of this population. Fourth, data is subject to internal monitoring and data quality checks performed within the CPRD (69). Finally, the CPRD contains long-term follow-up information on patient, enabling the study of long-term outcomes.

Study estimates were imprecise with wide confidence intervals, likely due to limited statistical power from small sample sizes in drug subclasses. Further, due to sample restrictions, analyses were limited to subsets of drug subclasses and outcomes. Our study may be subject to a level of exposure and outcome misclassification. Primary care data is indicative of prescriptions issued to patients but we are unable to confirm prescription dispensation or use. Although the CPRD is widely generalisable to the UK, prescriptions in primary care data for certain members of society, such as private patients or prisoners, may not be included. In this study, all patients received prescriptions before clinical confirmation of pregnancy. Therefore some ‘unexposed’ offspring may experience early intrauterine exposure before clinical confirmation of pregnancy. Hence, we may only assess the intentional treatment decision effects, not maternal prescriptive drug exposure effects on fetomaternal outcomes. Some literature indicates drug substance-specific adverse neonatal and maternal outcomes. We were unable to analyse the drug substance level due to low counts and sample size limitations. Our analysis estimated some OR of large effect; however, we note these may be implausible and occur due to low counts for certain outcomes and exposures. This highlights the need for further investigation in a larger dataset.

The CPRD contains complex patterns of missingness. Some patients may be misclassified due to missing information from their clinical record - for example, any diagnosis entered as free text would be excluded from our extract. Additionally, patients with greater health risk are monitored more rigorously than those with lower risk and have more complete records. This bias may vary with disease (21).

Therefore, our sample may not be reflective of the wider population. Within our study we generated and clinically validated lists of potential Read codes, medical codes, ICD-10 codes and OPCS codes that related to the conditions and variables of interest. Where possible we obtained externally validated lists to improve consistency of definitions between studies and aid comparability. Yet there is no standard, universal definition of disease diagnosis. In addition, data linkages are not available for all of the CPRD. For example, ONS mortality data is only available for deaths registered in England. Therefore, analyses requiring linkages have a limited sample size and findings may not generalise as well.

### Clinical implications

This study provides an update to the literature regarding the associations between maternal treatment discontinuation for hypertension and hypothyroidism and differential maternal and neonatal outcomes. We note the study is observational, thus cannot establish a causal relationship. Additionally, this study is subject to limitations such as precision and missing data, therefore our findings should be interpreted with caution alongside with evidence obtained from different study designs within the literature. We found evidence to suggest that discontinuation of thyroid hormone treatment was associated with reduced odds of miscarriage. This finding challenges some existing literature which suggests that thyroid hormone treatment reduces miscarriage rates. Further, our results indicated limited evidence linking the discontinuation of calcium-channel blockers and renin-angiotensin system drugs to specific outcomes. Discontinuation of many drug subclasses within this study did not show differential effects in this study. Clinicians should carefully evaluate the necessity of these medications during pregnancy and base decisions on a broader range of evidence and individual patient circumstances, considering both potential benefits and risks of continuing or discontinuing treatment.

### Research implications

It has been suggested in the literature that dose adjustment may be beneficial during pregnancy. For example, it has been proposed the dosage of thyroid hormone should be reassessed in line with BMI to effectively improve fetomaternal outcomes (57). Dosage optimisation was beyond the scope of our current study but remains a direction for future research. In addition, due to sample limitations and missing data we were unable to assess associations between maternal treatment discontinuation and many outcomes outlined within our protocol paper (33). Another possible direction for future research could be to implement this analysis in an alternate dataset of similar population where these variables may have lower levels of missingness, for example the Swedish medical birth register (58).

## Conclusions

In conclusion, our results suggest that large associations between many drug subclasses and adverse offspring outcomes are unlikely. We note our study is observational and limited to prescribing data, thus we cannot prove causal links between drug exposure during pregnancy and maternal or neonatal outcomes. However, evidence from this study may be used in conjunction with other studies to guide clinical decision-making for the treatment of chronic hypertension and hypothyroidism in pregnancy in similar populations.

## Data availability

Information on how to access CPRD data is available here: https://www.cprd.com/how-access-cprd-data.

## Acknowledgements

The Pregnancy register was derived using primary care data only. To recover information about “unknown” pregnancy outcomes, as defined in the Pregnancy register, and differentiate between conflicting or temporally overlapping pregnancy episodes I applied a proxy pregnancy-register algorithm to linked HES data. This algorithm was an adaptation of work by Dr Harriet Forbes and Dr Paul Madley-Dowd.

## Author contributions

CSB, VMW and NMD conceptualised the study. CSB acquired funding, curated the data, performed the formal analysis, created data visualisations and wrote the original draft. CB gave verification of medical codes and thresholds as indicated. VWM, CB, GDS and NMD supervised the study and reviewed and edited the article.

