## supplementary for "Assessing the benefits and risks to mothers and offspring of continuing treatment for maternal hypertension and hypothyroidism: an observational cohort study in the UK Clinical Practice Research Datalink"

### Supplementary materials

#### Supplementary methods

##### Additional details for the variable derivation

An external Read code list of ethnicities was obtained and regrouped into 5 wider categories: Asian, Black, Mixed, Other, White. The most commonly reported ethnicity within the mothers’ record was identified as their ethnicity within our study. If incidences of ethnicities were tied, we prioritised Mixed, Other Asian, Black then White (1).

Maternal BMI measures were restricted to those taken after the age of 16, and at most two years prior to the start of pregnancy, for each pregnancy identifier. Values less than 13kg/m^2^ and greater that 70kg/m^2^ were deemed implausible thus were excluded. We also calculated BMI for mothers with height and weight present. Any recorded height greater than 2.14m or below 1.3m was excluded and the median value was taken across multiple measures. Measures of weight were restricted to those after the age of 16 and prior to the start of pregnancy. Any recorded measures of weight less than 20kg or greater than 100kg were excluded. Thus, maternal BMI was defined as the nearest BMI measure to the start of pregnancy if there were multiple recorded values within the defined period.

Parity was calculated at the pregnancy identifier level and defined to be the number of outcomes determined to be a “delivery” (e.g. Live birth, Stillbirth, Live and Stillbirth, Delivery based on a third trimester pregnancy and Delivery based on a late pregnancy) within the Pregnancy register prior to the current pregnancy.

Where there were multiple differing delivery methods either per pregnancy identifier, per baby patient identifier or per pregnancy identifier per birth order, the final delivery method was determined as follows: prioritise caesarean section records, vacuum or forceps records, breech birth records, then “Other, unspecified" delivery.

#### Supplementary figures

Supplementary figure 1: A flowchart describing the variable derivation approach.

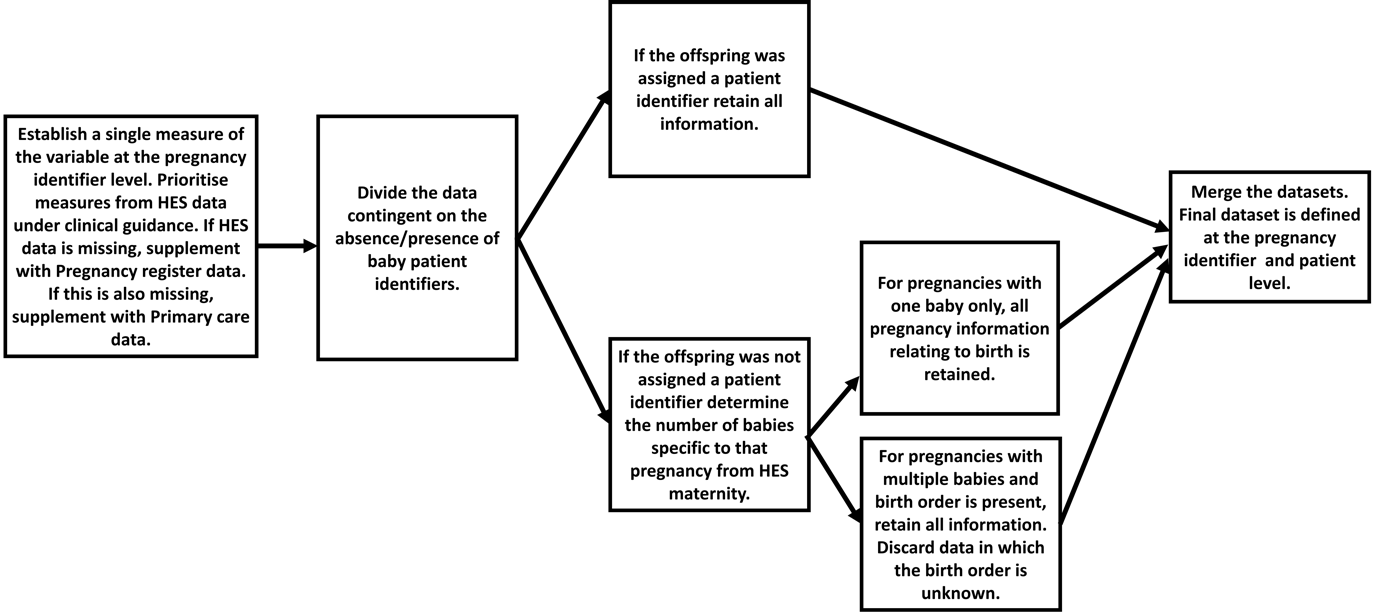

Supplementary figure 2: A comparative histogram demonstrating the propensity score distributions of pregnancies in which antihypertensive treatment was continued and discontinued, for pregnancies that reached a minimum gestational age of 22 weeks, and all gestational ages.
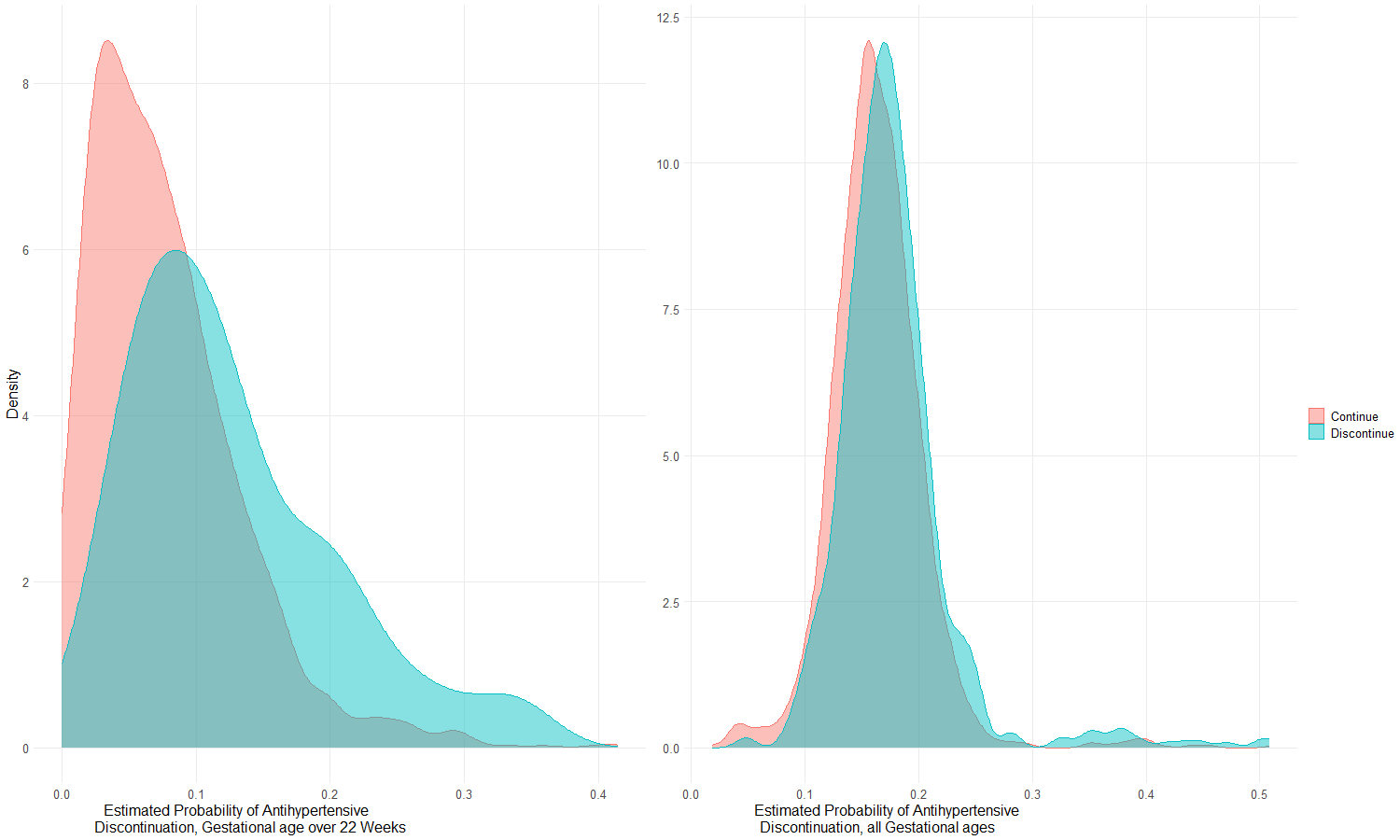

Supplementary figure 3: A comparative histogram demonstrating the propensity score distributions of pregnancies in which hypothyroid treatment was continued and discontinued, for pregnancies that reached a minimum gestational age of 22 weeks, and all pregnancies.

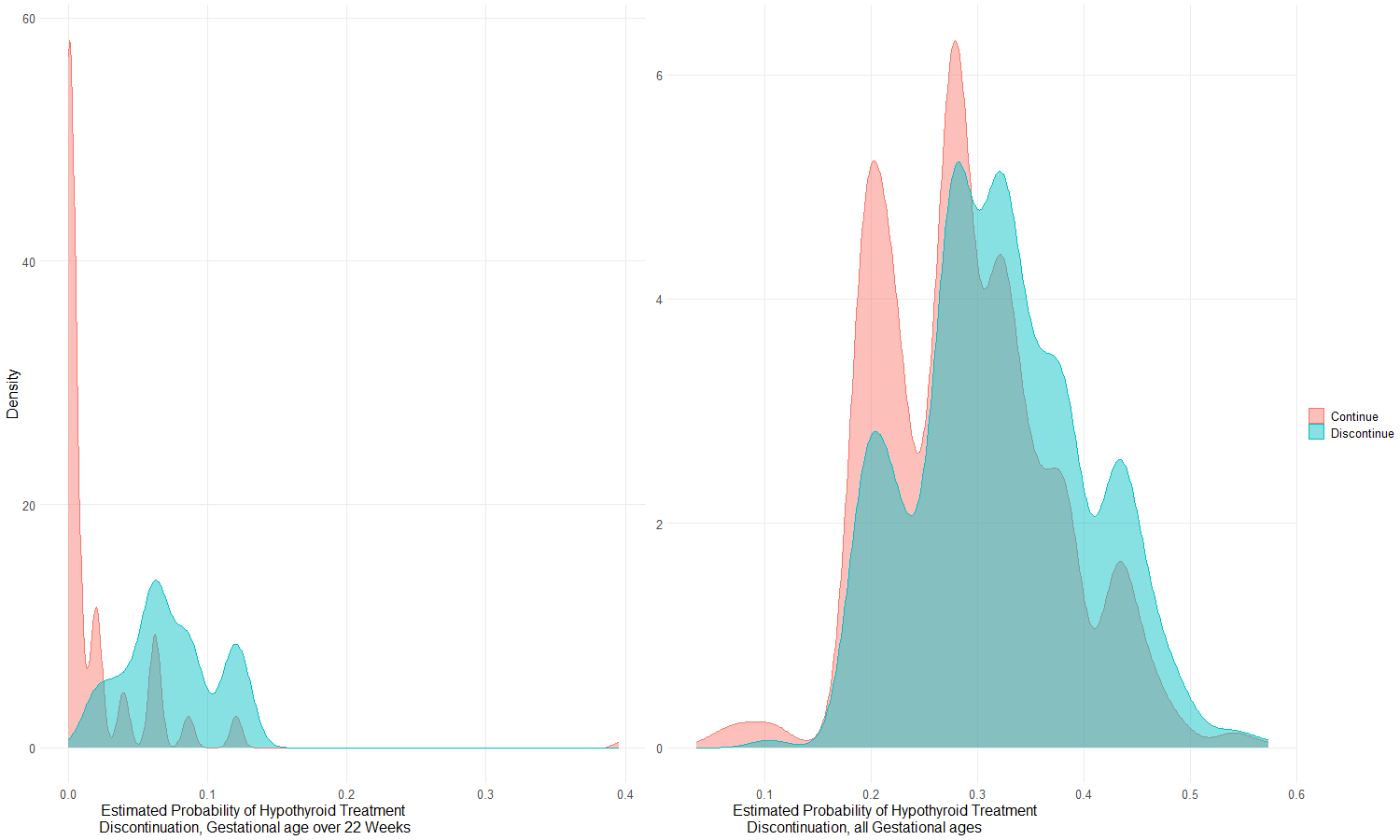

Supplementary figure 4: Forest plots demonstrating the estimated association between maternal discontinuation of treatment for antihypertensives, adjusted for propensity score. The odds ratio (OR) and 95% confidence interval (CI) has been calculated for binary outcomes (left), the mean difference and 95% CI has been calculated for continuous measures.

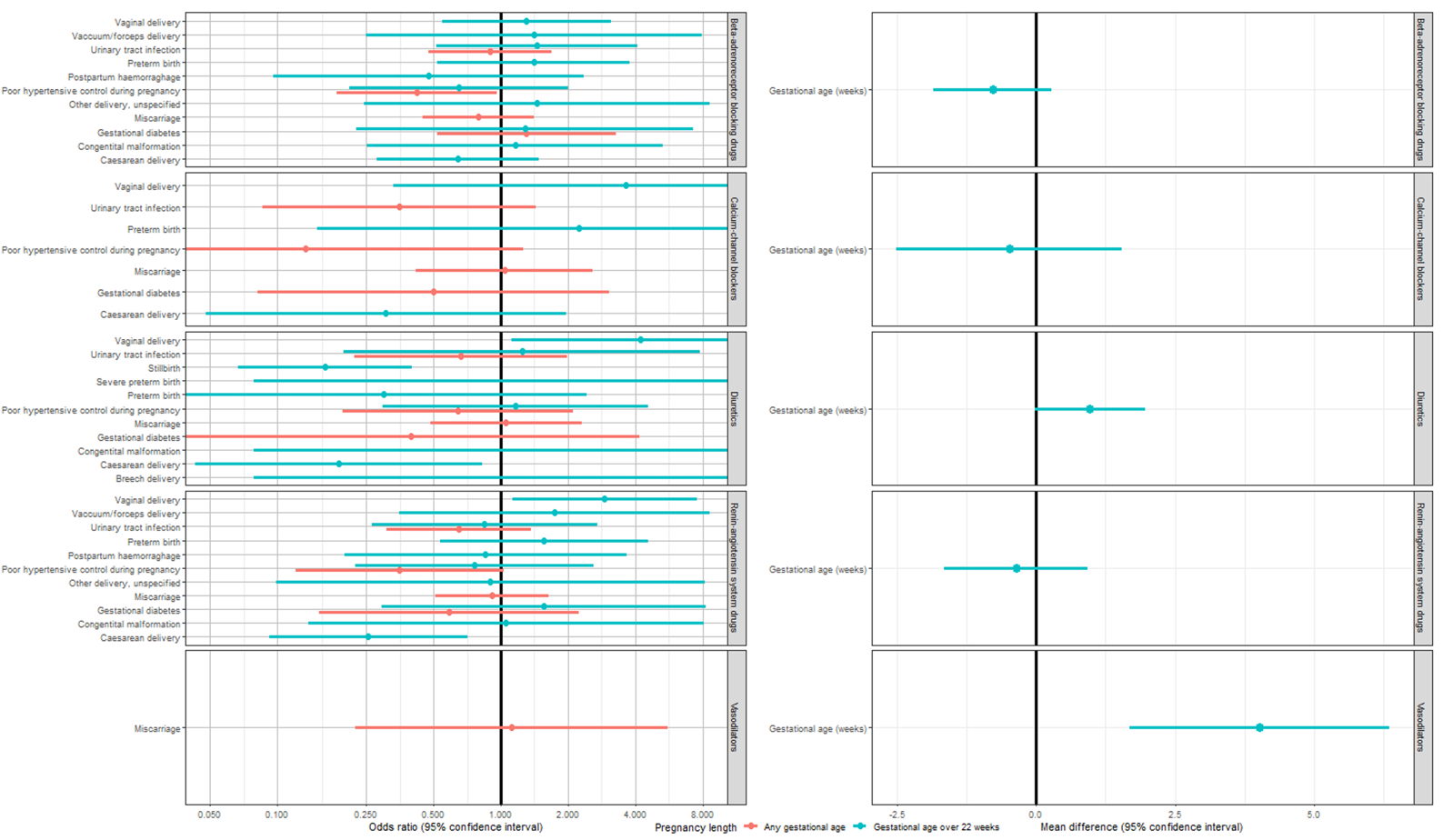

Supplementary figure 5: Forest plots demonstrating the estimated association between maternal discontinuation of treatment for treatment of hypothyroidism, adjusted for propensity score. The odds ratio (OR) and 95% confidence interval (CI) has been calculated for binary outcomes (left), the mean difference and 95% CI has been calculated for continuous measures.

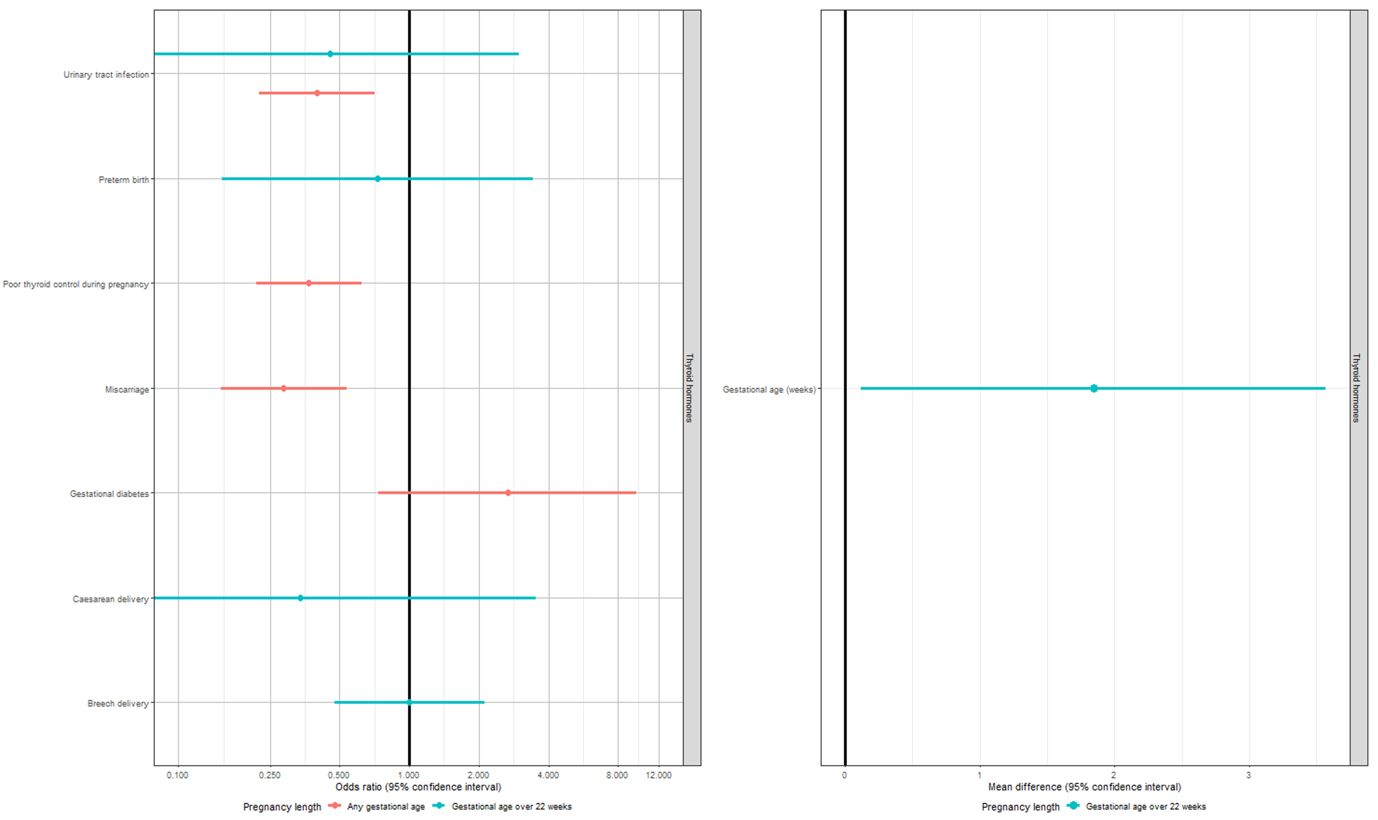

Supplementary figure 6: Forest plots demonstrating the estimated association between maternal discontinuation of treatment for antihypertensives using trimmed propensity scores. The odds ratio (OR) and 95% confidence interval (CI) has been calculated for binary outcomes (left), the mean difference and 95% CI has been calculated for continuous measures.

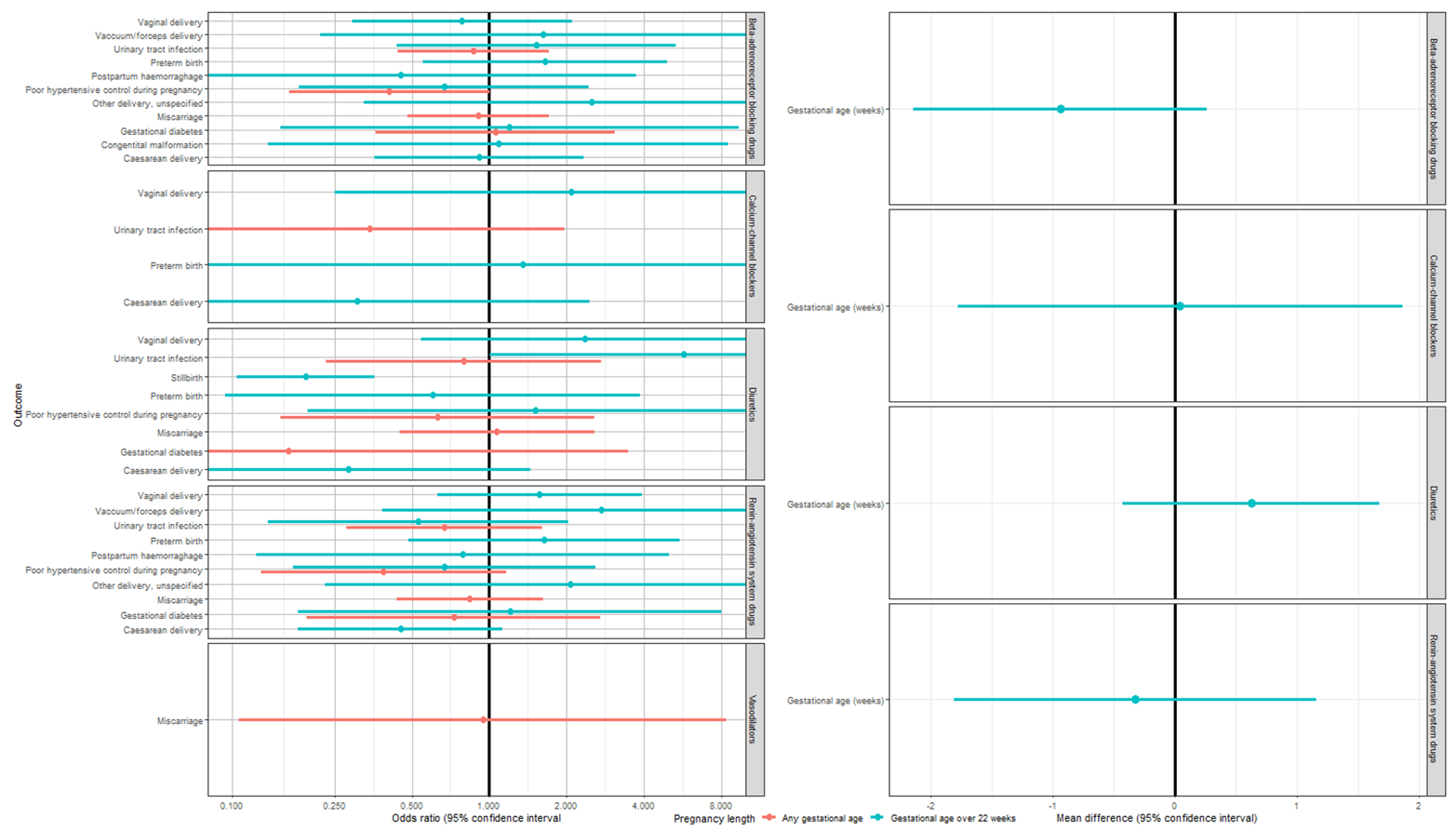

Supplementary figure 7: Forest plots demonstrating the estimated association between maternal discontinuation of treatment for treatment of hypothyroidism using trimmed propensity scores. The odds ratio (OR) and 95% confidence interval (CI) has been calculated for binary outcomes (left), the mean difference and 95% CI has been calculated for continuous measures.

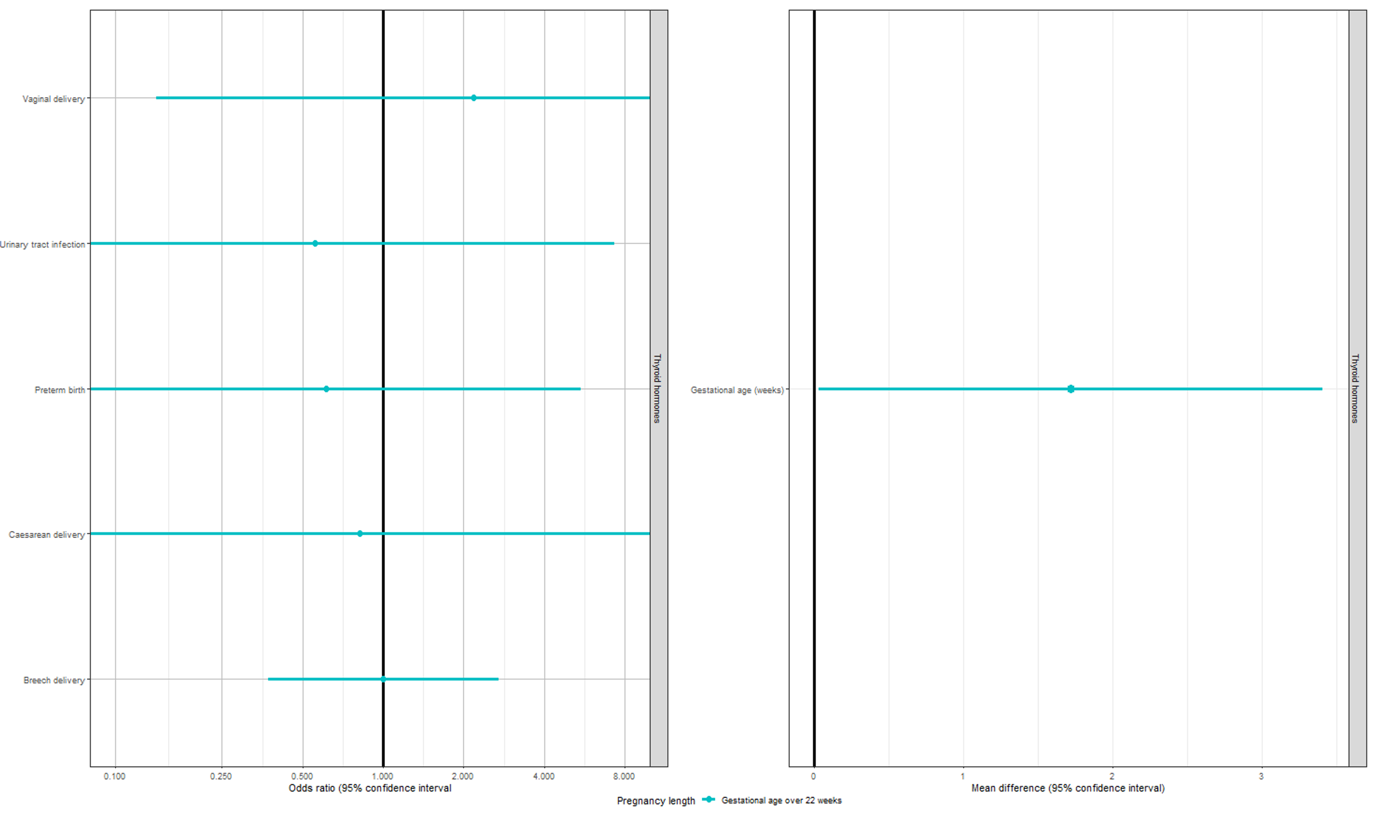

#### Supplementary tables

Supplementary table 1: A table describing the datasets used to derive study outcomes and covariates.

| **Variable** | **Dataset(s)** | **Code type** |
| --- | --- | --- |
| Live or still birth | Primary care data, Pregnancy register, HES maternity, HES procedures, HES diagnosis | Defined variable, medical codes, ICD-10 |
| Method of delivery | Primary care, HES diagnosis, HES maternity, HES procedure | Defined variable, medical codes, ICD-10, OPCS |
| Apgar score | Primary care data, HES diagnostic, HES hospital | Medical codes, ICD-10 |
| Sex of baby | Primary care data, HES maternity | Defined variable |
| Gestational age | Pregnancy register, HES maternity, HES procedures, HES episode | Defined variable |
| Maternal age | Pregnancy register, primary care | Derived variable |
| UTI | Primary care data, HES diagnosis, HES hospital diagnosis | Medical codes, ICD-10 |
| Preterm birth | Pregnancy register, primary care, HES maternity | Derived from gestational age, medical codes |
| Postpartum haemorrhage | Primary care, HES diagnosis, HES hospital diagnosis | Medical codes, ICD-10 |
| Poor hypertensive control during pregnancy | Primary care, HES diagnosis, HES hospital diagnosis | Medical codes, ICD-10 |
| Miscarriage | Primary care, pregnancy register | Medical codes, defined variable, |
| Poor thyroid control during pregnancy | Primary care, HES diagnosis, HES hospital diagnosis | Medical codes, ICD-10 |
| Gestational diabetes | Primary care, HES diagnosis, HES hospital diagnosis | Medical codes, ICD-10 |
| Severe preterm birth | Pregnancy register, primary care, HES maternity | Derived from gestational age, medical codes |
| Congenital malformation | Pregnancy register, primary care, HES maternity | Derived from gestational age, medical codes |
| Historical incidence of postpartum haemorrhage | Primary care | Medical codes |
| Historical miscarriage | Primary care | Medical codes |
| Historical stillbirth | Primary care | Medical codes |
| Historical preterm birth | Primary care | Medical codes |
| Multiple birth | Pregnancy register | Defined variable |
| Smoking status prior to pregnancy | Primary care | Medical codes, product codes |
| Teetotal prior to pregnancy | Primary care | Medical codes |
| Relationship status | Primary care | Defined variable |
| Maternal age | Pregnancy register | Defined variable |
| Maternal body mass index (BMI, kg/m^2^), up to two years prior to pregnancy | Primary care | Medical codes |
| Maternal ethnicity | Primary care, HES patient, HES episodes | Medical codes, defined variable |
| Parity | Pregnancy register | Derived variable |

*BMI; body mass index, CPRD; Clinical Practice Research Datalink, HES; Hospital episode statistics, ICD-10; International Classification of Diseases 10th Revision, OPCS; Office of Population Censuses and Surveys Classification of Interventions and Procedures, UTI; urinary tract infection*
